# Saliva testing as a means to monitor therapeutic lithium levels in patients with psychiatric disorders: identification of clinical and environmental covariates, and their incorporation into a prediction model

**DOI:** 10.1101/2021.03.23.21253841

**Authors:** Georgia M. Parkin, Michael J. McCarthy, Soe H. Thein, Hillary L. Piccerillo, Nisha Warikoo, Douglas A. Granger, Elizabeth A. Thomas

**Author notes:** Correspondence: Georgia M. Parkin, Ph.D., Department of Epidemiology, University of California, Irvine Irvine, CA.

## Abstract

The narrow therapeutic window of lithium medications necessitates frequent serum monitoring, which can be expensive and inconvenient for the patient. The use of saliva as a biofluid may have advantages over blood, as it is non-invasive, easier to collect, requires less processing, and can be collected without the need for trained personnel. This study investigated the utility of saliva as a longitudinal means of monitoring lithium levels. We measured lithium levels using Inductively-Coupled Plasma Optical Emission Spectrometry (ICP-OES) in n=171 saliva samples collected via the passive drool method, from a multi-center cohort consisting of n=75 patients with bipolar disorder or other psychiatric conditions. We found that saliva and serum levels of lithium were highly correlated (unadjusted Spearman r=0.74, *p*<0.0001) and that consideration of daily lithium dose, dosing schedule, cotinine-confirmed smoking status and diabetes status could improve this relationship (adjusted Spearman r=0.77, *p*<0.0001). Using this adjusted intersubject equation to predict an individual’s serum lithium levels from their salivary lithium value, we observed a strong linear correlation between the predicted vs. actual serum lithium levels r=0.70; P<0.0001). Longitudinal samples were collected from patients for up to 18 months from the initial assessment. The saliva/serum ratios across these multiple visits were highly stable for most patients. Variability in the saliva/serum ratios across observations was found to be significantly associated with age. Using the intrasubject saliva/serum ratio from a single prior observation was not better than using the interpatient linear regression equation at predicting the serum lithium levels. However, the using the mean intrasubject ratio calculated from three prior observations could robustly predict serum levels with a predicted vs. actual serum correlation of r=0.90 (*p*<0.0001). These findings strongly suggest that saliva could be used for lithium monitoring and open the door for the development and implementation of a point-of-care salivary lithium device that could be used at home or in the clinic. We propose that the use of saliva will dramatically improve treatment opportunities for patients with mood disorders.

| **Significant Outcomes** |  |
| --- | --- |
| 1 | Salivary lithium measurements can accurately predict serum lithium levels, however optimal monitoring must account for confounding clinical and lifestyle factors. |
| 2 | Use of a patient's average intrasubject saliva/serum lithium ratio from three preceding appointments shows improved predictive power and may be particularly advantageous to combat variability present in older patients. |
| 3 | Implementation of salivary lithium monitoring, including a home-based collection design, would lead to a personalized monitoring approach for patients taking lithium medications, and would improve treatment opportunities for patients with mood disorders |
| **Limitations** |  |
| 1 | This study included a limited number of samples from patients within the in higher range of serum lithium levels. More research will be required to validate an intersubject prediction model for patients nearing the level of lithium toxicity. |
| 2 | This study included patients who may have been non-compliant or missed their scheduled dose; the prediction of serum lithium levels from saliva may be improved by only considering patients who are stable and compliant. |

**Data availability statement:** Anonymized summary data will be shared by reasonable formal request from qualified researchers, subject to a data sharing agreement and in compliance with the requirements of the funding bodies and institutions.

## Introduction

Lithium was identified as a treatment for bipolar disorder (BD) in 1949 and remains a first line of treatment for both mania and depression^[1,2]^ with particular benefit in reducing suicidality. While highly effective, the use of lithium is limited by its narrow therapeutic range (0.5-1.2 mmol/L), and common occurrence of adverse effects, such as gastrointestinal discomfort, tremors, drowsiness, disorientation, and renal and thyroid dysfunction. These side effects are commonly dose related, but can occur even at therapeutic doses^[3-7]^. At higher serum concentrations, the potential for side effects and risk of toxicity increases further^[8]^. The presence of side effects may contribute to poor outcomes and medication non-adherence^[9]^. For these reasons, lithium requires frequent monitoring of blood serum levels. International guidelines recommend that patients on lithium medication undergo lithium level monitoring at least every 3-6 months, and even more frequently after dosage change, in case of relapse, or upon symptoms of intoxication^[10,11]^.

Serum lithium monitoring requires attendance at a clinical laboratory and the collection of a blood sample, which can be uncomfortable, expensive and inconvenient for patients. The results may vary across time due to a variety of factors including lifestyle and/or diet-promoted changes in body composition and lithium clearance, and alterations in lithium levels may go undetected for months^[12]^ Lithium is stored in the adrenal glands, bone, lymph nodes and pituitary gland, and eliminated predominantly via the kidneys^[3]^. As such, factors affecting renal function, such as age, dehydration, caffeine and heavy or chronic alcohol intake, sodium balance and hemodynamics, serum creatinine levels and body mass index (BMI) also have the potential to impact on serum lithium levels^[12-15]^. Hence, providing an effective and more convenient means for testing would allow for more frequent lithium monitoring, optimal dosing, and improved patient outcomes with lithium medications.

Previous studies dating back 30 years have suggested the use of saliva as an alternative to blood monitoring in humans^[16-18]^. Several of these studies demonstrated significant correlations between serum lithium and salivary lithium in adult patients being treated with lithium carbonate^[19-22]^, although, other studies found only weaker associations^[23-26]^. These past studies suffered from inconsistencies in saliva collection and storage procedures, variations in saliva processing methods, differences in the lithium detection method and overall provided little information regarding patient demographic and clinical features^[16]^. Nonetheless, saliva sampling provides enormous potential clinical utility as a non-invasive, cost-effective approach for lithium level monitoring that can be carried out in any setting. Indeed, the potential for patients to collect saliva samples at home would reduce the need for trained laboratory personnel, and allow patients to collect a sample at a time that is not only convenient for them, but can also be better coordinated with their daily dose, timing of other medication and/or food intake. In addition, saliva collection is safer than blood draws, as it does not require the use of a needle, which can increase the transmission of blood-borne pathogens.

In this study, we explored the relationships between lithium levels in saliva and serum in a multicenter cohort of patients taking lithium medications and confirmed a strong correlation between saliva and blood serum lithium levels. We further identified a number of clinical and lifestyle factors that affect the lithium levels in these two biological fluids, and demonstrate the potential for saliva to represent an alternative non-invasive biofluid for lithium monitoring.

## Material and Methods

### Human Subjects

This study was approved by Institutional Review Boards at the University of California Irvine (UCI), University of California San Diego (UCSD) and VA San Diego Healthcare System (VASDHS), in accordance with the requirements of the Code of Federal Regulations on the Protection of Human Subjects. Patients were recruited from the UCI Medical Center (UCIMC), and VASDHS. Subjects (n=75) were recruited through the physical and online posting of a flyer, and through recommendation from their psychiatrist. Individuals were eligible to participate if they were age 3 or older, were currently taking lithium medication, and were receiving either inpatient or outpatient psychiatric treatment. All individuals provided informed, written consent prior to sample collection. Subjects were given the opportunity to participate in this study up to 8 times, providing up to 8 longitudinal, time-matched saliva/blood samples, spaced at least one month apart. After recruitment, two subjects were excluded due to non-compliance with medications and a third due to a history of stage 3 kidney disease.

### Medical data collection

Demographic, medical and blood chemistry results were collected on the day of sample collection, through the electronic medical record system. Data collected included age, sex, smoking status, medical diagnoses, lithium formulation and dose, and any concurrent psychiatric medications. Where available, blood chemistry data included the serum levels of sodium, potassium, creatinine, thyroid stimulating hormone, and white blood cell count.

### Sample collection

Saliva samples were collected using the gold standard passive drool method^[27]^. All donors were asked to refrain from smoking, eating, drinking, or oral hygiene procedures for at least 1 hour prior to sample collection. Roughly 1-2 milliliters of unstimulated whole saliva was obtained. Samples were stored at −20 °C within an hour of collection, and remained at −20°C for up to several weeks, and were then stored at −80 °C. Following saliva collection, subjects were either accompanied, or directed to, the medical center pathology clinic, where serum lithium levels were measured using standard hospital procedures.

### Lithium analysis

At the time of saliva sample analysis, occurring after at least 24 hours storage at −80 °C, saliva samples were thawed and centrifuged (10,000 g; 5 min; 4 °C) to remove insoluble material and cellular debris. The resulting supernatant was collected and used for all assays. Salivary lithium levels were measured via Inductively-Coupled Plasma Optical Emission Spectrometry (ICP-OES) (Avio200; Perkin Elmer, Waltham, MA, USA) using the radial viewing window. Prior to analysis, the ICP-OES was aligned with 0.1% manganese. For ICP-OES analysis, samples were diluted 1:20 in 2% HNO_3_; output was normalized to the internal standard, 0.1% yttrium, and compared to a lithium standard curve. All yttrium recovery values fell between 89% and 106%. The second lithium standard curve point (1000 ppm lithium), as well as a HNO_3_ blank, were re-run after every 10 samples, to ensure consistent analysis parameters and the absence of sample carry-over, respectively. In addition, prior to the analysis of study samples, we first tested whether the levels of a spiked-in, known amount of lithium varied according to saliva collection method (passive drool vs. swab), saliva processing method (centrifuged vs. uncentrifuged), storage conditions (−20°C, 4°C, room temperature), or the number of freeze-thaw cycles. We found that lithium levels did not vary according to processing method, storage temperature up to 24 hours, nor the number of freeze-thaws up to three (Suppl. Figure 1). However, we did observe that lithium recovery was slightly lower in swab collection devices compared to passive drool (p=0.05) (Suppl Figure 1). As passive drool is considered the gold standard method approved for all salivary analytes^[27]^, this method of saliva collection was used for this study.

### Total protein and cotinine assays

Salivary total protein was measured via Pierce™ BCA Protein Assay (Thermo Fisher Scientific, Waltham, MA) at a 1:10 dilution in dH_2_O. Salivary cotinine was assayed in duplicate using a commercially-available ELISA kit (Salimetrics LLC, Carlsbad, CA) following the manufacturer’s protocol. The assay has a test volume of 20 µL, range of standards from 0.8 to 200 ng/mL, and lower limit of detection of 0.15 ng/mL.

### Determination and validation of lithium prediction models

For the development of a prediction model, data instances were randomly assigned to either a determination cohort, which contained 75% of all data points (n=127), or a preliminary validation cohort, which contained the remaining 25% of data points (n=42). Randomization was conducted using the Microsoft Excel random number generator, and splitting the resulting randomly ordered data sets into two consecutive groups. The determination cohort was used to determine the prediction model, the validation cohort was used to validate the prediction model.

### Statistical Analysis

Analyses were conducted with GraphPad Prism version 8.4.2 for Windows (GraphPad Software, La Jolla, CA, USA). Analyses which corrected for covariates were conducted using IBM® SPSS® Statistics version 25 for Windows (IBM Corp., NY, USA). Outliers were identified using the ROUT outlier test (Q=5%), resulting in the exclusion of two outlier saliva lithium measurements. The measurements of salivary lithium, serum lithium levels, and saliva/serum ratios were not normally distributed (Skewness= 0.99, 0.31 and 0.94, for saliva, serum and ratio, respectively). Consequently, all data were analyzed using nonparametric analyses, with the exception of prediction model determination and validation. As outcome variable normality is not a required assumption for linear regression modelling, modelling was analyzed without data transformation, and using Pearson correlations. Non-parametric Spearman correlations which corrected for covariates were conducted by ranking continuous data, followed by multivariate linear regression. For the correlation comparison of lithium levels and blood chemistry data, as well as the comparison of lithium levels and other instance-specific variables such as daily lithium dose, data points were included from all available visits. Comparisons between lithium levels and static continuous data, such as age and BMI, were conducted using all subjects’ first time point data only.

## Results

### Patient characteristics and study variables

In this study, we collected 171 saliva samples from n=75 patients across two recruitment sites, UCIMC and VASDHS. After recruitment, two subjects were excluded due to non-compliance with medications and a third due to a history of stage 3 kidney disease. Demographic characteristics for the cohorts recruited from UCIMC and VASDHS are outlined in Table 1. The age range for patients was 14 – 76 years, with 74.6% of patients being male. All patients recruited from VASDHS had a diagnosis of bipolar disorder, whereas 74% of patients from UCIMC had a diagnosis of bipolar disorder and 15% had a diagnosis of either schizophrenia, schizoaffective disorder, major depressive disorder or generalized anxiety disorder. Cohorts differed significantly in sex distribution, age and BMI, but not salivary or serum lithium levels (Table 1). Salivary lithium levels were two to three-fold higher compared to those measured in serum, and these ratios were slightly different between sites (2.38 vs. 2.75, for UCIMC and VASDHS, respectively; Table 1).

**Table 1.** Demographic and disease data for subjects included in this study (median, range).

|  | All subjects<br>(n=75) | Subjects by site |  |  |
| --- | --- | --- | --- | --- |
|  |  | UCIMC<br>(n=38) | VASDHS<br>(n=37) | *p* |
| **Sex [M/F]** | 56/19 | 22/16 | 34/3 | 0.002 |
| **Diagnosis [BD/Scz/MD]** | 65/5/5 | 28/5/5 | 37/0/0 | 0.001 |
| **Age [years]<sup>#</sup>** | 35.2, 14.5-76.0 | 28.6, 14.5-58.0 | 52.0, 26.0-76.0 | <0.0001 |
| **BMI<sup>#</sup>** | 29.1, 20.1-48.0 | 27.8, 20.1-39.0 | 31.0, 22.0-48.0 | 0.02 |
| **Daily Lithium dose [mg]** | 900, 300-2400 | 900, 300-2400 | 900, 300-1800 | 0.95 |
| **Saliva lithium levels**<br>**[mmol/L]** | 1.36, 0.23-3.54 | 1.34, 0.29-3.03 | 1.42, 0.23-3.54 | 0.32 |
| **Serum lithium levels**<br>**[mmol/L]** | 0.58, 0.10-1.20 | 0.56, 0.10-1.14 | 0.61, 0.13-1.20 | 0.92 |
| **Saliva/Serum lithium ratio** | 2.44, 1.09-5.50 | 2.38, 1.09-5.50 | 2.75, 1.12-5.23 | 0.005 |

Sex and diagnosis differences were analyzed with a Fisher's Exact Test, with diagnoses grouped into 'bipolar disorder' or 'other'; Differences in continuous measures between sites were analyzed with the Mann-Whitney U test; BD, Bipolar Disorder; Scz, Schizophrenia or Schizoaffective disorder; MD, Mood Disorder (Major Depressive Disorder or Anxiety). <sup>#</sup>Variables compared using data from the first appointment only, all other measures above used data from multiple appointments.

Both salivary and serum lithium levels were positively correlated with daily lithium dose and serum lithium levels were significantly affected by dosing schedule (once or twice a day) (Table 2). A diagnosis of type 2 diabetes, and smoking, the latter confirmed through measurement of salivary cotinine levels, were associated with variations in the saliva/serum lithium ratios (Table 2). No significant effects of age, sex, BMI, lithium formulation (immediate release vs. sustained release) or total protein levels (a measure of dehydration) were found to be associated with lithium levels in saliva, serum, nor the saliva/serum ratios (Table 2).

**Table 2.** Factors affecting serum and saliva lithium measures.

| Variable | Continuous measures<br>(Spearman rho, p) |  |  |
| --- | --- | --- | --- |
|  | Salivary Li | Serum Li | Ratio |
| Age<sup>#</sup> | 0.12, 0.32 | 0.10, 0.40 | 0.09, 0.47 |
| BMI<sup>#</sup> | 0.01, 0.92 | -0.02, 0.85 | -0.06, 0.62 |
| Daily lithium dose | **0.26, 0.001** | **0.40, <0.0001** | -0.16, 0.06 |
| Salivary total protein | -0.04, 0.70 |  | -0.12, 0.28 |
|  | Non-continuous measures<br>(U, p) |  |  |
|  | Salivary Li | Serum Li | Ratio |
| Sex | 1916.0, 0.18 | 1898.0, 0.49 | 1768.0, 0.33 |
| Smoking status | 3007.0, 0.52 | 2760.0, 0.50 | **2203.0, 0.03** |
| Diabetes status | 1175.0, 0.36 | 1138.0, 0.40 | **832.5, 0.04** |
| Lithium formulation<sup>1</sup> | 3041.0, 0.76 | 2404, 0.08 | 2353, 0.14 |
| Lithium once vs twice per day | 2427.0, 0.42 | **1945, 0.02** | 1987, 0.08 |

<sup>1</sup> (immediate vs. sustained release). Continuous measures were analyzed by Spearman correlation analysis. Categorical measures were analyzed using Kruskal-Wallis non-parametric tests. <sup>#</sup>Variables compared using data from the first appointment only, all other measures above used data from multiple appointments.

### Correlation between saliva and serum lithium levels

Linear regression analysis across all samples revealed a significant correlation between saliva and serum lithium levels (unadjusted Spearman rho r=0.74, b=0.26, p<0.0001) (Figure 1A).

**Figure 1.**
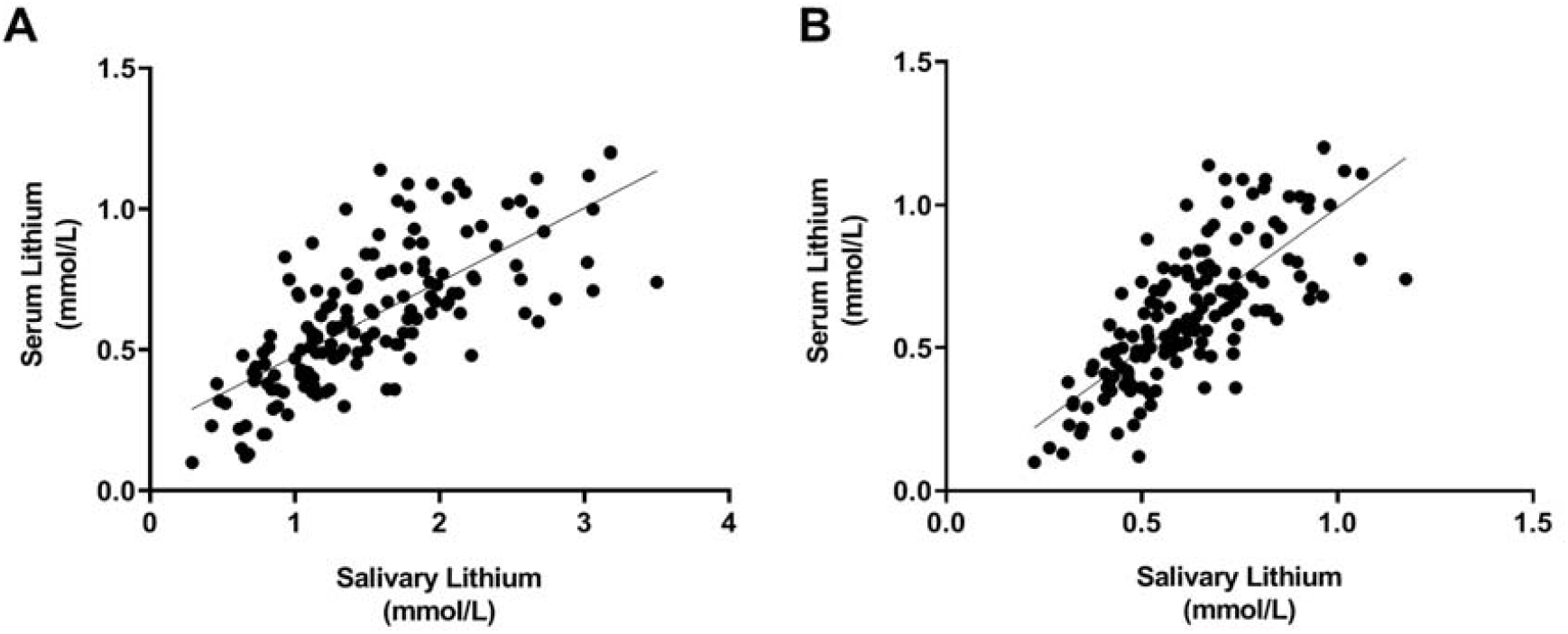
Correlations between saliva and serum lithium levels, unadjusted (A), and adjusted for relevant covariables, (B). Unadjusted correlation was determined by Spearman correlation analysis (r=0.74; p<0.0001). Adjusted correlation (Spearman rho=0.77, p<.0001) was determined by multivariate linear regression analysis including adjustment for daily lithium dose, type 2 diabetes and smoking.

Subsequently, multivariate regression including stepwise consideration of the significant variables from Table 2, as well as those that differed between recruitment sites (age, sex and BMI) revealed significant effects of daily lithium dose (*p*<.0001), a diagnosis of type 2 diabetes (*p*=0.010), and smoking (*p*=0.03) on the association between salivary lithium and serum lithium. Incorporation of these covariates into the equation modestly improved the saliva-to-serum correlation (Spearman rho=0.77, p<.0001; Figure 1B). Significantly, this multivariate linear regression had a slope of b=1.0, suggesting a 1:1 adjusted relationship between salivary and serum lithium levels (Figure 1B).

### Associations between blood chemistry and lithium levels

Blood chemistry data, including serum sodium, potassium and creatinine levels, were available for a subset of the samples (n=80-85) enabling investigation into whether these factors affect lithium levels in serum or saliva or the saliva/serum ratio. Serum and salivary lithium levels were found to be significantly associated with serum potassium levels (Table 3); this correlation remained statistically significant after adjusting for recruitment site, daily lithium dose, dosing schedule, diabetes status, and smoking status (Table 3). In contrast, the saliva/serum lithium ratio was not correlated with any measure of blood chemistry (Table 3).

**Table 3.**
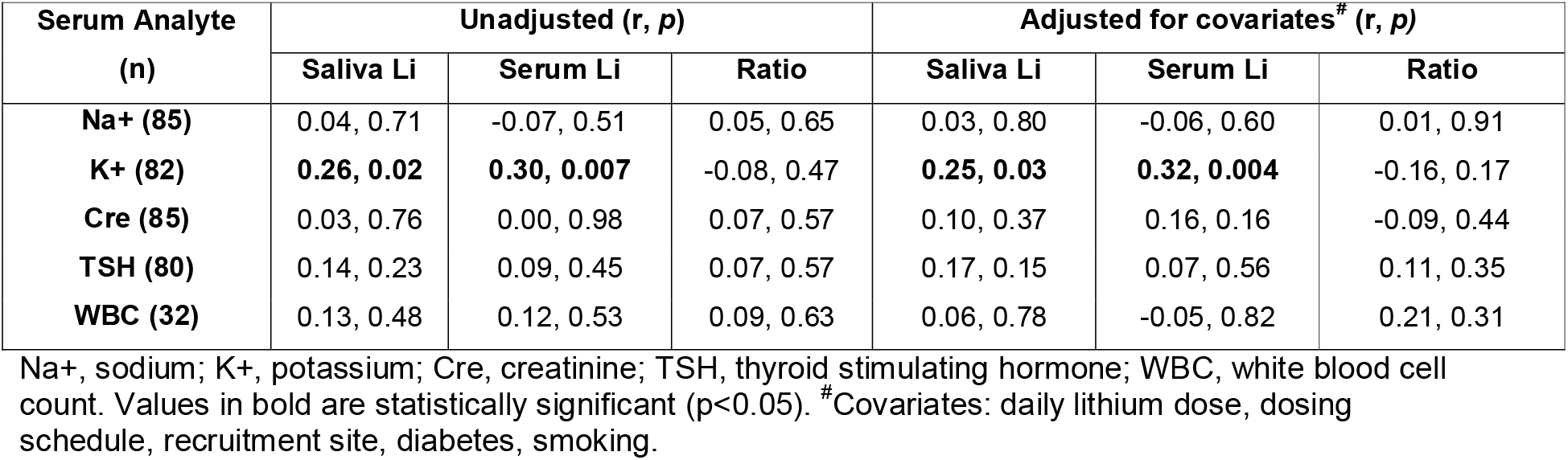
Relationships between biofluid lithium levels and blood chemistry

### Predicting serum lithium levels from salivary levels across patients

We next explored how well salivary lithium levels could predict serum lithium levels across all saliva samples, which were split into a “Determination” cohort (n=127) and a preliminary “Validation” cohort (n=42), as described in Methods. Multivariate linear regression carried out on the Determination cohort, including stepwise correction for daily lithium dose, type 2 diabetes, smoking and recruitment site as covariables, revealed similar results to that observed with the entire cohort, with the exception of smoking status, which was no longer significant. The adjusted saliva-to-serum correlation coefficient value of the Determination cohort was identical to that observed with the whole cohort (Prediction model Pearson r=0.77; Figure 2A).

**Figure 2.**
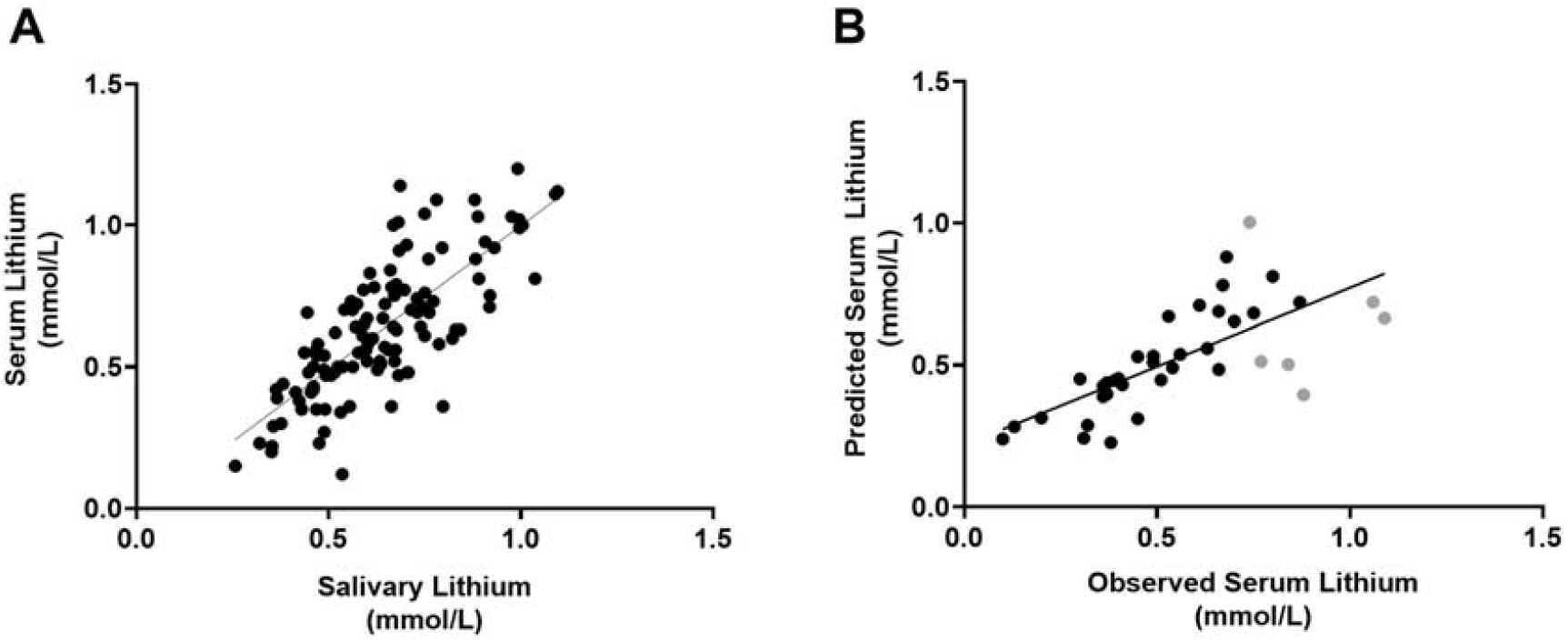
Relationship between salivary and serum lithium levels in the Determination cohort (A) and the relationship between predicted and observed serum lithium levels in the Validation cohort (B). The correlation between the saliva and serum lithium values in panel A were adjusted for daily lithium dose, diabetes status and recruitment site (Pearson r= 0.77, *p*<0.0001). The correlation between the observed and predicted serum lithium levels using the validation cohort equation from 2A was significant at a Pearson r= 0.70, *p*<0.0001. Predicted serum lithium levels that differed from observed levels by more than 0.2 mmol/L are highlighted in grey in panel B.

Next, we tested the prediction model on the validation cohort (n=38) using the following linear equations, solving for Y (serum lithium levels): Y= 0.166 + 0.255*(Saliva lithium) −0.056*(1 if VASDHS, 0 if UCIMC) + 0.145* (1 if diabetic, 0 if not).

The salivary lithium levels predicted serum lithium levels with a Pearson correlation rho value of 0.70 and slope (b) of 0.89. Of the 38 samples included in this model, 6 (16%) of the predicted serum levels differed from observed levels by more than 0.2 mmol/L (shown in grey in Figure 2B).

### Predicting serum lithium levels from saliva levels within patients

Over half of the patients in our study (n=40) provided samples on multiple visits, which allowed for the prediction of serum lithium levels using the patient’s own previous salivary lithium data. We used the saliva/serum ratio from each observation in combination with their salivary lithium level from the patient’s subsequent appointment to predict the serum level for that second appointment. The saliva/serum ratios for each appointment for these patients is shown in Suppl. Table 1. Using this approach, we found the predicted vs. observed serum lithium levels to be significantly correlated (Pearson r=0.71; p<0.0001; Figure 3A), even when patients had a dose change from one appointment to the next. Patients attending four or more appointments allowed us to compare additional within-subject data to determine if the prediction of serum lithium level would be improved by calculating the value based on the average of the saliva/serum ratios from the first three visits. The use of subjects’ average within-subject saliva/serum lithium ratio from three prior observations, dramatically improved the ability to predict serum levels from saliva on a fourth observation of the same subject. (Pearson r=0.90, b=1.3, *p*<0.0001), albeit with a small sample size (n=16) (Figure 3B). Using this method, 75% of samples showed <0.1 mmol/L difference in the predicted vs. observed serum lithium level and only two samples out of 16 showed a difference > 0.2 mmol/L.

**Figure 3.**
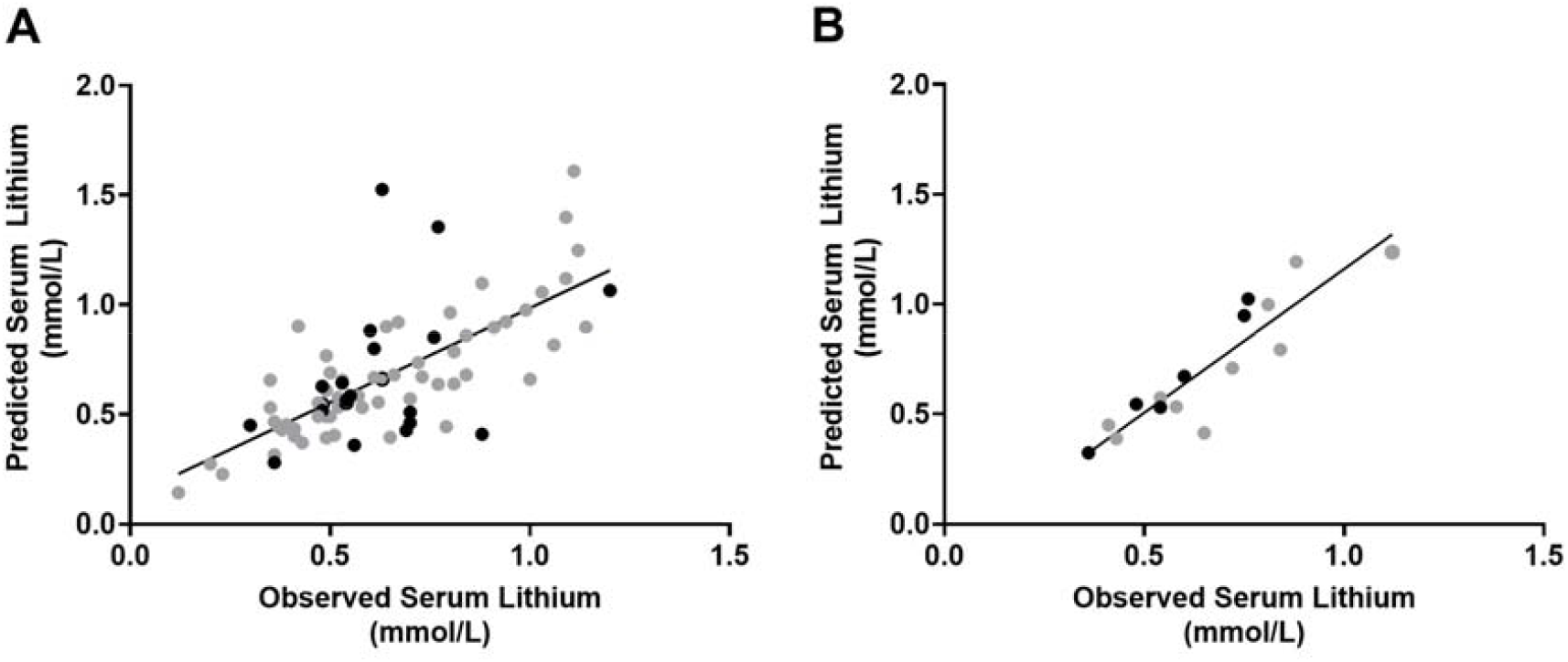
Comparisons of predicted vs. actual lithium levels, based on a single prior within-subject saliva/serum lithium ratio (A) and an average of three prior within-subject saliva/serum ratios (B). In panel A, the predicted vs. observed serum lithium levels were significantly correlated (Pearson r=0.71; p<0.0001), however this correlation improved dramatically when an average of three ratios were used to predict the 4th appointment (r=0.90; p<0.0001). Overall, patients above the age of 55 showed the most variability in saliva/serum lithium ratio levels (black data points in A, B). Patients were included in Figure 3A if they attended two or more appointments, and Figure 3B if they attended four or more appointments.

Overall, the within-subject prediction of serum lithium levels based on salivary lithium levels from a single preceding appointment alone was not better than that calculated across different patients. This is likely due to the high variation in saliva/serum ratios observed from one appointment to the next for some patients (Figure 4; Suppl. Table 1), although it is clear that many patients show stable saliva/serum ratios across several months, and even up to 18 months (Figure 4; Suppl. Table 1). As lithium pharmacokinetics may be affected by age-related physiological changes, and because of the very large age range of our patients (14-76 years), we assessed the effect of age on the variability in saliva/serum ratios across any two appointments. Indeed, there was a significant correlation between age and the variation of the saliva/serum ratio across observations (Spearman r=0.40; p=0.0001; Suppl Figure 2), an effect which was no longer significant after including only patients <55 years of age. Furthermore, when comparing within-subject linear regressions from patients <55 years compared to ≥55 years (Figure 3A, grey vs. black data points), the regression of patients <55 years passed a test for homoscedascity, whereas the regression of patients ≥55 years did not (*p*=0.03). This increase in residual variation was overcome by using the average saliva/serum lithium ratio from the preceding 3 visits (both *ps*>0.05; Figure 3B grey vs black data points).

**Figure 4.**
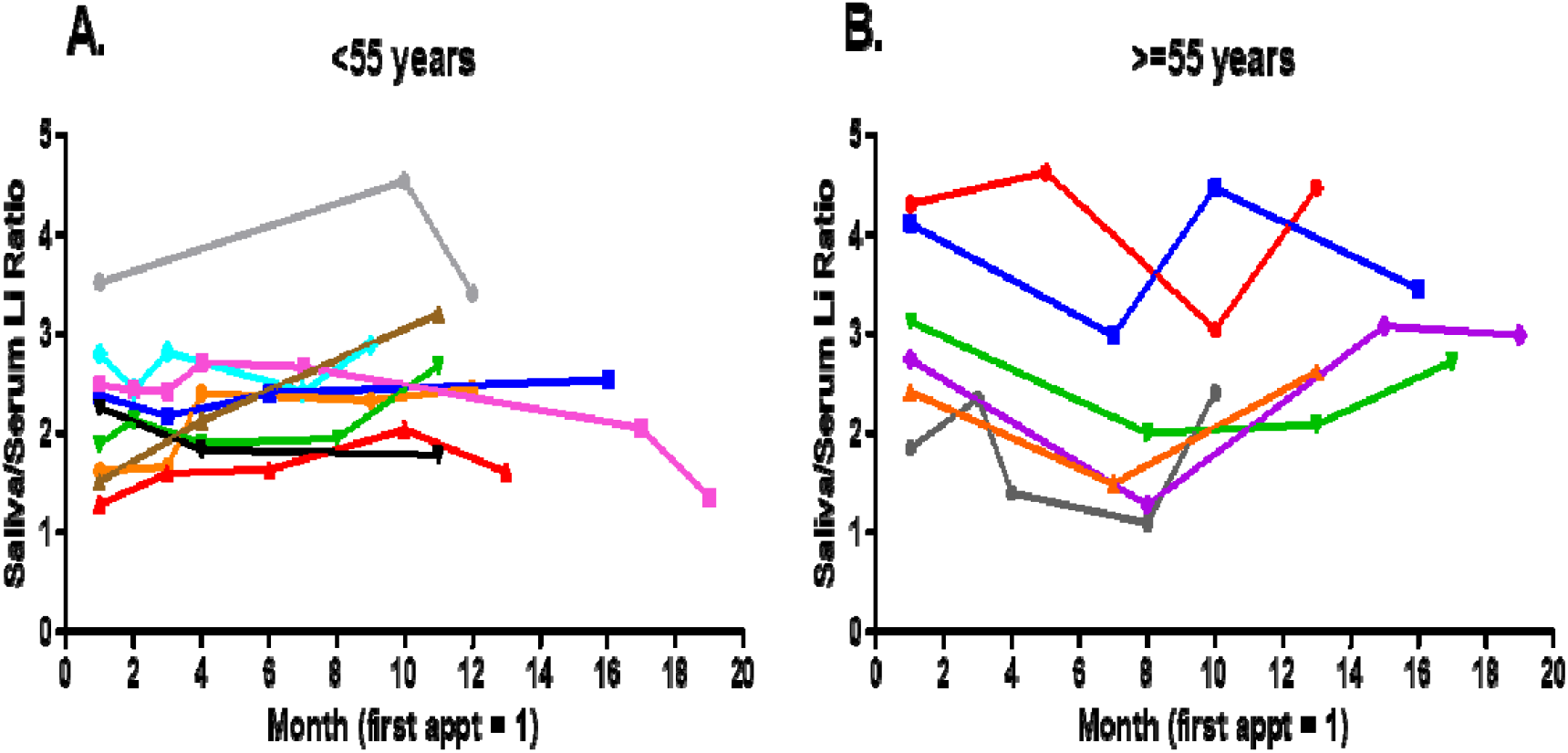
Saliva/serum ratios for individual patients across multiple visits in patients younger than 55 years (A) and equal or greater than 55 years (B). Patients’ first study appointment is represented by month 1, followed by every subsequent appointment, spaced by months since the first appointment. For clarity, only data from patients attending visits spanning 10 months or greater from their initial appointment are shown. Each color represents an individual patient. Saliva/serum ratios for all patients attending at least two appointments are shown in Suppl Table 1.

## Discussion

In this study, we found that salivary lithium levels are highly correlated with serum lithium levels across 169 matched saliva-serum samples from 75 subjects recruited from two different sites, strongly supporting the suitability for use of saliva for monitoring of lithium levels in patients taking lithium medications. The use of saliva for drug monitoring has been reported previously. In fact, a large body of work dating back >30 years has suggested that salivary lithium might be useful for patient monitoring of lithium levels, with several previous studies demonstrating good correlation between saliva and serum lithium values (i.e. between 0.50 to 0.88) ^[16,28-33]^. However, in past studies, the sample sizes were small, saliva collection techniques varied, older methods for lithium quantification were used and medical and lifestyle variables were not included^[16]^. Furthermore, few past studies have investigated the longitudinal within-patient association between salivary and serum lithium levels^[34]^ as we have in the current study. With improved methodology, inclusion of important covariates, and a larger sample derived from a multi-center cohort, we have conducted the largest and most integrative study to date supporting the utility of saliva as a relevant biofluid for therapeutic lithium monitoring.

In this study, we have shown that incorporation of daily lithium dose, dose regimen, smoking status and diabetes can improve the predictive power of saliva for serum lithium levels. Our covariate-adjusted inter-subject model had similar predictive power to the use of intra-subject ratios from a patient’s single preceding appointment, but was less powerful than the use of an average intra-subject ratio from a patient’s preceding three appointments. In clinical practice, both measures have their own advantages. For example, the covariate-adjusted between-subject model could be used in the instance that a patient does not have three preceding, saliva/serum lithium ratio measurements.

Previous studies have shown that body weight, serum creatinine, serum potassium and age are correlated with lithium levels in serum^[35-38]^. In this study, we did not observe an association with BMI, nor creatinine, on serum lithium levels, which is consistent with some other studies^[37,39]^. We did find significant associations between serum potassium levels and both saliva and serum lithium levels, but not with saliva/serum lithium ratios. Nonetheless, consideration of serum potassium could be important given that potassium intake has been proposed to play a role in chronic kidney disease^[40]^, which is associated with diabetes, a condition we found to significantly affect the saliva-serum lithium correlation. Further, we found that the variability in saliva/serum lithium ratios across time was positively correlated with age. As glomerular filtration rate decreases with age^[15]^, older individuals may become more sensitive to lithium treatment. Indeed, blood lithium concentration appears to be higher in the elderly, after adjusting for dose, compared to younger patients, which could be due to an age-related reduction in renal function and increase in disease comorbidity, and more use of common medications that may interfere with lithium elimination such as hydrochlorothiazide, NSAIDS, and ACE inhibitors and angiotensin receptor antagonists^[14]^. In addition, elderly subjects are more likely to have multiple comorbidities which could have yet-undetermined effects on lithium distribution in the body. These previous studies may explain why patient saliva/serum lithium ratios were more variable in older patients, and together, suggest that within-subject lithium prediction might be limited to younger populations. However, and importantly, our data also suggests that for patients above the age of 55, use of their average saliva/serum lithium ratio from three preceding appointments, rather than one preceding appointment or the between-subject regression, may circumvent this variation.

Advantages to using saliva rather than blood are many, including the fact that it doesn’t require trained personnel to collect and can be carried out in any setting; these features could directly translate into more frequent and improved lithium monitoring for both outpatients and inpatients. For outpatients, saliva sampling could allow for at-home collection, which would allow patients to schedule lithium measurements around last dosing, food and other activities, without the need to arrange and attend a pathology clinic. As such, at-home sampling would more accurately predict an individual’s trough lithium level. This is important because prescription of lithium medications is based on the trough level in the blood, which is difficult to capture precisely in a clinical appointment. While we found no association between time lapsed from last dose to blood collection, and either salivary lithium, serum lithium or saliva/serum lithium ratios, previous research by Serdarevic et al. which compared salivary lithium and serum lithium correlations at 2- and 12-hours post-last dose, suggested that the saliva-serum correlation may be improved by collecting samples closer to the last dose^[32]^. Also, because of the known difference in lithium pharmacokinetics in saliva vs. serum [32,41], the timing of the last dose is of critical importance. For inpatients, saliva measures could also be used to better optimize the timing of trough levels, as saliva sampling would not be restricted to a phlebotomist’s schedule. Further, and as we observed in our study, inpatients are much more likely to agree to a saliva collection versus a blood draw, although this feature is not limited to inpatients, as many outpatients, especially those in a manic state, are also averse to blood draws.

Importantly, we found that lithium measurements were stable at 4 °C or room temperature for at least 24 hours. This feature would further facilitate a home-health care-based program that could involve saliva sample collection at home, followed by samples sent to a centralized lab for lithium measurements. The results could then be monitored and discussed remotely via a telemedicine encounter. Such a process could avoid costly office, and maybe even hospital, visits.

Our study is not without limitations. One limitation was that it included a limited number of samples from patients within the in higher range of serum lithium levels (≥1 mmol/L), and it is within this range that the between-subject prediction of serum lithium levels fell by greater than 0.2 mmol/L outside of the observed levels (see Figure 3B). Given the upper limit of toxicity for serum lithium is 1.5 mmol/L, predicting serum levels from saliva levels in patients taking high doses of lithium could be problematic when using the intersubject model with regards to potential toxic side effects. However, when serum levels were predicted using the average intrasubject ratio from three appointments, the predicted serum level was less than 0.1 mmol/L from the observed level for 75% of the patients, with two patients having predicted levels >0.2 mmol/L. This suggests accurate estimations of serum lithium using the average ratios, although future studies to better characterize saliva lithium levels at the upper boundaries of the therapeutic range and into the toxic range will be needed to more fully characterize the value of saliva testing of lithium before implementation into routine clinical practice. Additionally, the value of salivary measures would need to be calibrated to an individual, as there are currently no FDA standards for a stand-alone saliva measure.

Another limitation of our study was that some participants were non-compliant with their dose, or missed their scheduled dose. Because we wanted to maximize the power of our study, these patients were not excluded; however, give the significant effect of lithium daily dose on the saliva-serum relationship, we expect that improved results would be obtained using only those patients on stable medication and those who are compliant with their medications. Of note is that our prediction models included samples from some patients who changed their dose across subsequent visits, suggesting that a dose change or dose titration would not greatly affect the ability of saliva to predict the serum level. Nonetheless, the notion that the inter-subject model would be best applied to patients who are clinically stable, as well as medically compliant and might only require testing every 3-6 months, was also suggested in a previous study by Rosman and colleagues^[34]^. Additionally, given that biweekly lithium testing is commonly indicated shortly after starting the drug and/or after changing dose, additional examination of saliva testing should focus on subjects undergoing these changes to better characterize the sensitivity and saliva/serum ratio when lithium levels are dynamic.

Considering the overall literature, despite variations in methods of salivary lithium collection and quantification, the majority of previous studies have reported saliva/serum lithium correlations similar to that observed in our study^[28-33]^. This consistency in results across studies, as well as the stability in saliva/serum lithium ratios in most patients across time observed in our study, opens the door for the development and implementation of a point-of-care (POC) salivary lithium device. A saliva-based POC assay for lithium would have enormous value in the new era of personalized medicine, as it would lead to optimized treatment protocols for at-home collection as well as patient compliance in the clinic. In 2005, the FDA approved the use of a finger-prick in-office test for serum lithium levels^[42]^. Use of this POC serum test can already reduce the frequency and need for laboratory-conduced venepuncture, however, the kit is expensive, and has issues related to separation of blood components. Conversely, saliva requires less processing than blood, and it is therefore estimated that it would not be as expensive to operate. While such devices would require standard collection procedures, our studies suggest that sampling at room temperature did not affect lithium quantitation and that levels did not vary whether the samples were centrifuged or not (Suppl Figure 1). It is important to note, however, that not all potential contributors to saliva/serum lithium level variation have been investigated. It is therefore possible that use of a novel at-home device may be improved further through the incorporation of, or affected by the presence of, other as of yet un-identified variables. These could include factors such as sugar intake, water intake, caffeine and sleep patterns.

In summary, our findings show robust correlations between saliva lithium levels and those measured in serum, and that saliva lithium values can accurately predict serum lithium levels, especially when prior intrasubject data is considered. Implementation of salivary lithium monitoring, including a home-based collection design, would lead to a personalized monitoring approach for patients taking lithium medications, and would improve treatment opportunities for patients with mood disorders, although additional studies on the efficacy of salivary lithium measurements in a more controlled patient cohort would be needed. Additionally, a POC saliva assay would provide the basis for future technical innovations that could allow for lithium monitoring via point-of-care devices that could transmit data directly to the patient’s medical provider through cloud-based data storage or a smartphone. The implementation of such devices in saliva samples is already underway^[43]^.

## Data Availability

Anonymized summary data will be shared by reasonable formal request from qualified researchers, subject to a data sharing agreement and in compliance with the requirements of the funding bodies and institutions.

## Acknowledgements

This study was funded by the Elisabeth Severance Prentiss Foundation. The authors wish to thank Andrew Huang for excellent technical assistance.

