## Supplementary Data for "Saliva testing as a means to monitor therapeutic lithium levels in patients with psychiatric disorders: identification of clinical and environmental covariates, and their incorporation into a prediction model"

Supplementary Figure 1. The effect of sample treatment conditions on saliva lithium levels. Known lithium concentrations were spiked into saliva samples from five psychiatric patients, and tested for the impact of saliva collection method (passive drool vs. swab), processing (centrifuged or 'spun' vs non-centrifuged or 'unspun'), or storage for 24 hours at room temperature (RT), vs 4°C vs -20°C. All comparisons other than that of saliva collection method were conducted using the passive drool samples.

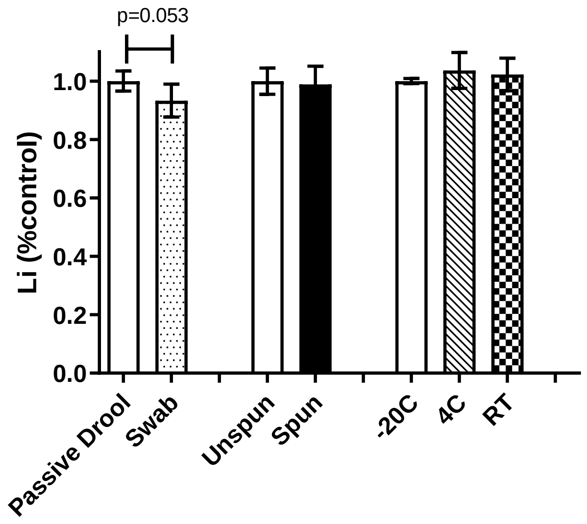


Supplementary Figure 2. Correlation of the variation in saliva/serum ratios across any two visits with age. Spearman correlation, r=0.397; p=0.0001.

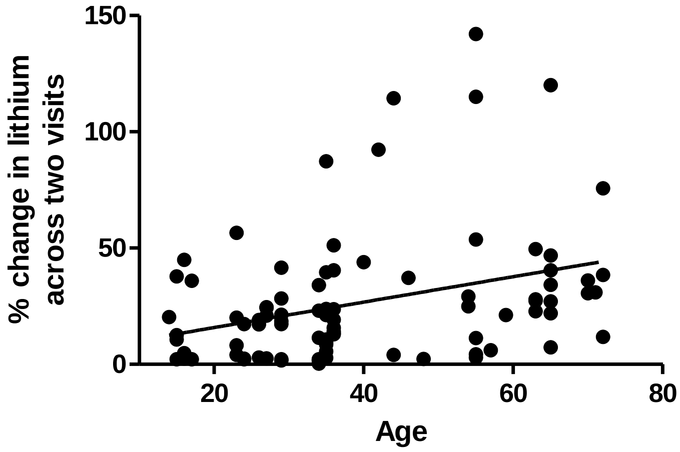

Supplementary Table 1. Summary of the saliva/serum ratios of patients attending multiple visits.

| **Record number** | **Lithium dose/day** | **Appt. 1** | **Appt. 2** | **Appt. 3** | **Appt. 4** | **Appt. 5** | **Appt. 6** | **Appt. 7** |
|---|---|---|---|---|---|---|---|---|
| **1** | 1200 | 2.14 | 1.77 | 1.74 | 1.77 | 1.39 | 1.79 | 1.46 |
| **2** | 1500 | 2.49 | 2.44 | 2.43 | 2.71 | 2.67 | 2.05 | 1.35 |
| **3** | 600 | 2.13 | 1.95 | n.c. | 1.87 | 2.93 | 2.34 | |
| **4** | 2400 | 1.62 | 1.66 | 2.41 | 2.33 | 2.44 | | |
| **5** | 600 | 2.80 | 2.44 | 2.82 | 2.42 | 2.88 | | |
| **6** | 1200 | 1.28 | 1.59 | 1.63 | 2.03 | 1.61 | | |
| **7** | 750 | 1.89 | 2.13 | 1.90 | 1.94 | 2.68 | | |
| **8#** | 300 | 1.85 | 2.35 | 1.40 | 1.09 | 2.40 | | |
| **9** | 900 | 1.81 | 2.13 | 2.08 | 2.13 | | | |
| **10** | 1200 | 2.76 | 3.02 | 3.10 | 1.88 | | | |
| **11#** | 900 | 2.75 | 1.27 | 3.08 | 2.99 | | | |
| **12#** | 600 | 4.31 | 4.63 | 3.04 | 4.47 | | | |
| **13#** | 900 | 1.39 | 2.99 | 2.65 | 2.76 | | | |
| **14#** | 900 | 4.11 | 2.99 | 4.47 | 3.45 | | | |
| **15#** | 900 | 2.42 | 1.49 | 2.62 | 2.93 | | | |
| **16#** | 900 | 3.12 | 2.00 | 2.09 | 2.72 | | | |
| **17** | 1200 | 2.38 | 2.17 | 2.40 | 2.54 | | | |
| **18** | 300 | 2.93 | 2.87 | 3.90 | | | | |
| **19** | 1500 | 2.27 | 1.83 | 1.78 | | | | |
| **20** | 900 | 1.21 | 2.60 | 2.70 | | | | |
| **21** | 1200 | 1.51 | 2.12 | 3.20 | | | | |
| **22** | 450 | 5.23 | n.c. | 3.05 | | | | |
| **23** | 1500 | 3.82 | 4.73 | 3.73 | | | | |
| **24** | 1350 | 3.52 | 4.54 | 3.41 | | | | |
| **25** | 600 | 2.33 | 2.65 | | | | | |
| **26** | 1800 | 2.42 | 3.84 | | | | | |
| **27** | 600 | 3.23 | 3.31 | | | | | |
| **28** | 900 | 2.61 | 3.23 | | | | | |
| **29** | 1800 | 4.69 | 5.50 | | | | | |
| **30** | 600 | 2.63 | 3.16 | | | | | |
| **31** | 1500 | 1.41 | 2.71 | | | | | |
| **32#** | 300 | 2.23 | 2.92 | | | | | |
| **33#** | 600 | 2.54 | 3.07 | | | | | |
| **34#** | 600 | 1.97 | 2.09 | | | | | |
| **35** | 1500 | 2.17 | 2.22 | | | | | |
| **36#** | 600 | 4.72 | 3.40 | | | | | |
| **37** | 1200 | 4.00 | 2.24 | | | | | |
| **38** | 1800 | 2.24 | 3.07 | | | | | |
| **39** | 900 | 1.85 | 3.46 | | | | | |

n.c. Non-compliant with blood test. #Patient was 55 years of age or older
